# Progesterone and hCG in expectant management success in tubal ectopic pregnancy: retrospective single-centre cohort study

**DOI:** 10.64898/2026.08.05.26359789

**Authors:** Ayesha Karim Ahmad, Mark Pandrich, Aditi Naik, Aria Astruc, Kirsten Lafferty, Nishel Mohan Shah, Dede Ofili-Yebovi

## Abstract

**Background:** Early access to pregnancy assessment units now detects many tubal ectopic pregnancies (TEP) at a stage when they could resolve spontaneously, creating a management dilemma.

**Methods:** We performed a hypothesis-generating exploratory analysis in a retrospective study to assess whether serum progesterone (P4) levels in women with TEP are associated with management outcome.

**Results:** Ninety-one cases of TEP managed in a single centre over three years were analysed. Receiver operating characteristic (ROC) curve analysis was used to explore serum levels of progesterone (P4), first human chorionic gonadotropin (hCG) and peak hCG (alone and in combination) in relation with successful completion of expectant management. Decision-tree analysis using first hCG and P4 was additionally performed to explore clinical sequential risk stratification. 23% (n=21) successfully completed expectant management. P4 concentrations in the expectant management group (median 3 nmol/L, IQR 2.00–8.50) were significantly lower than in those requiring surgical or medical management (median 17 nmol/L, IQR 5.75– 29.25; p=0.0002). Area under the ROC curve (AUC) values for P4, log10 first hCG, log10 peak hCG and P4 with log10 first hCG were 0.766, 0.814, 0.811 and 0.835, respectively, for predicting successful expectant management. However, hCG was not significantly outperformed. Nonetheless, Youden optimised thresholds for hCG and P4 are reported, alongside decision-tree analysis that identified sequential first hCG and P4 thresholds associated with successful expectant management.

**Conclusion:** Lower P4 levels are associated with successful expectant management of TEP but they do not outperform hCG either alone or as an adjunctive marker.

**Lay summary:** Early pregnancy scans now identify many small, symptom-free fallopian tube pregnancies; human chorionic gonadotrophin (hCG) is a pregnancy hormone that can predict if these pregnancies will resolve naturally but it has its limitations. This study looked at 91 women with a fallopian tube pregnancy. It compared another important hormone called progesterone to see if, when used with hCG, its levels are associated with natural resolution. This study found that hCG was the most useful test, and progesterone did not provide any additional benefit.

## Introduction

Ectopic pregnancy affects 11 in 1000 pregnancies in the United Kingdom. Although the outcomes of ectopic pregnancy have improved significantly, it remains a common early-pregnancy related cause of death. The MBRRACE (Mothers and babies: reducing risk through audits and confidential enquiries across the UK) review of early pregnancy deaths from 2021-2022, showed ectopic pregnancy was the leading direct cause of maternal death and this risk has nearly doubled compared to 2018-2020 (Cantwell et al., 2011, Collaboration, 2022). Improved access to early pregnancy scanning, has led to earlier detection of tubal ectopic pregnancy (TEP), and more frequently in asymptomatic women (Kirk et al., 2006, Rajkhowa et al., 2000). Many of these will resolve spontaneously but determining which will regress and which will progress to life-threatening complications remains a clinical challenge.

Current NICE guidelines use human chorionic gonadotrophin (hCG) levels alongside ultrasound findings and clinical presentation to determine suitability of expectant management of TEP, recommending this approach when hCG levels are <1000 IU/L and considering it when <1500 IU/L (NICE, 2023). However, there continues to be wide variation in practice in the published literature. Low hCG levels of <175 IU/L have been shown to be highly predictive of successful expectant management (Elson et al., 2004). However, intermediate hCG levels are not as easy to interpret. Studies using hCG levels of <1500 IU/L to triage women as suitable find that a significant proportion (20-30%) fail expectant management (Elson et al., 2004, Mavrelos et al., 2013). A threshold of <1000 IU/L similarly misclassifies 12% of cases (Trio et al., 1995). Other studies suggest that selected cases of TEP with high hCG levels (3000-5000 IU/L) may be successfully managed expectantly (Helmy et al., 2015, Rodrigues et al., 2012). However, relatively few TEPs are managed in this way, and the evidence base remains limited. In general, as hCG levels rise, the success of expectant management decreases but not always in a linear fashion. Consequently, there is greater diagnostic uncertainty at higher hCG levels and failure rates of up to 21% have been reported when expectant management is applied with initial hCG level >1500 IU/L (Elson et al., 2004). In our study a >1500 IU/L cut-off hCG exclusion criteria were applied for expectant management of TEP based on local protocol and NICE guidance (NICE, 2023).

Progesterone (P4) is an essential hormone in pregnancy, the level of which increases with gestational age (Stjernholm, 2012, Chernecky and Berger, 2013, Arck et al., 2007). Low levels of P4 have been associated with pregnancy failure or non-viable pregnancy (Mol et al., 1998, Verhaegen et al., 2012, Abdelazim et al., 2013), and may be used to predict the likelihood of complete miscarriage for women presenting with a threatened miscarriage (Tan et al., 2020). Consequently, P4 levels may be assessed at time of presentation in these cases. Currently, the interpretation and significance of P4 levels in the context of TEP remains controversial and is not included in standard clinical guidance.

P4 concentrations ranging from <3.2ng/ml (10.9nmol/L) to 45ng/ml (153 nmol/L) have been associated with adverse early pregnancy outcomes (Mol et al., 1998, Verhaegen et al., 2012, Tan et al., 2020, Abdelazim et al., 2013). However, studies have failed to demonstrate if P4 levels can be used to differentiate between TEP and non-viable intrauterine pregnancies (Mol et al., 1998, Verhaegen et al., 2012, Cabar et al., 2008), with some demonstrating that it can be used as a sensitive but not specific marker (Dart et al., 2002). Nonetheless, P4 levels may provide additional information once the diagnosis of TEP has been established. Tubal rupture is more likely to occur in ectopic pregnancy with ongoing embryonic growth. Conversely, failing ectopic pregnancies have lower P4 levels and are more likely to resolve. In contrast, P4 levels in a normally developing pregnancy exceeds both (Ku et al., 2018). The aim of this study was to explore whether serum P4 levels, alone or as an adjunct to hCG, are associated with management outcome and specifically successful expectant management. Preliminary findings were presented by the authors as a conference abstract at the RCOG World Congress 2023. The current manuscript represents a full and expanded report of the study (Pandrich et al., 2023).

## Methods

This was a retrospective cohort study at a single London teaching hospital with N=91 participants. In our institution, all women presenting with pain and/or bleeding in the first trimester are referred to our early pregnancy assessment unit (EPAU) for a high resolution transvaginal pelvic ultrasound using Voluson E6 and S10 Expert GE Medical Systems machines. All women diagnosed with a TEP over a 3-year period (October 2017 to December 2020) were eligible for inclusion. Given the retrospective design, no *a priori* sample size or power calculation was undertaken. The study population comprised all consecutive eligible cases within the defined study period, yielding 61 participants in the initial combined surgical/medical management group and 33 in the expectant management group (Figure 1). This pragmatic approach limited statistical power, particularly for subgroup analyses. On that basis, successful expectant management was compared to the combined surgical/medical management group for the utility of P4.

**Figure 1.**
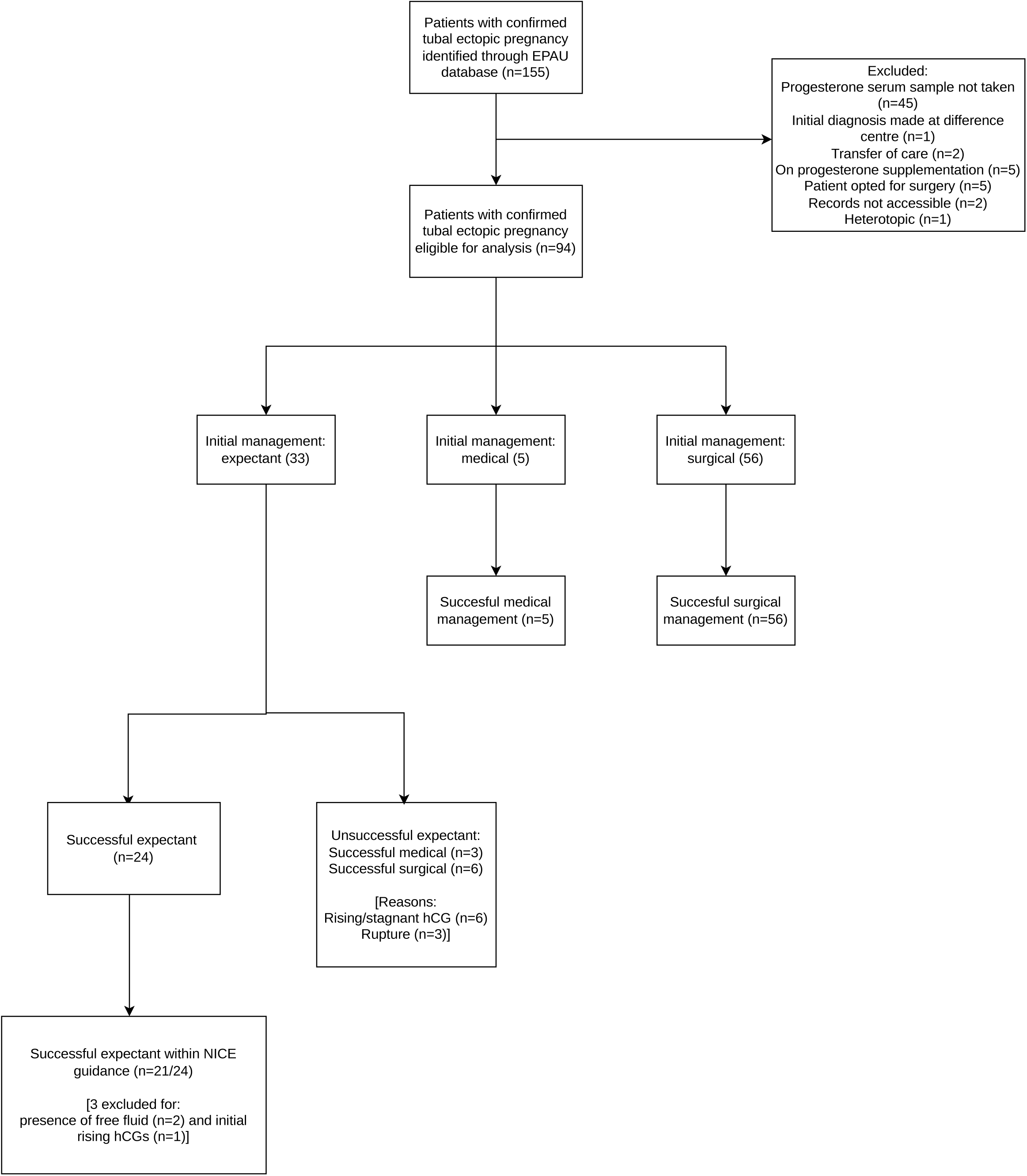
A Flow diagram of patient inclusion and management. Patients were identified using electronic patient records during the study period. Of the total (N=155) records, patients where serum P4 levels were not available and if they did not meet inclusion criteria for expectant, surgical or medical management of ectopic pregnancy were excluded from analysis. The remainder were analysed (N=91) and separated into expectant or surgical/medical combined management groups based on NICE criteria.

Expectant management was offered to women with TEP as per local guidance between 2017 to 2019, and then NICE (2019) guidance adopted thereafter. Specifically this was offered to clinically stable women with minimal symptoms, with a non-viable TEP (no visible heartbeat) on ultrasound, without significant hematoperitoneum and where serum hCG was <1000 IU/L and considered where serum hCG <1500 IU/L. Peripheral blood was collected in Serum Separator Tubes (SST2 tubes, Becton Dickinson, Wokingham, UK) from participants at first attendance to hospital. Serum P4 and first hCG were assessed at presentation. Timepoints for additional hCG measurements varied between participants. Peak HCG was defined as the highest recorded hCG. Serum hCG and P4 were measured on the Abbot Architect and Alinity platforms (Abbott, Berkshire, UK) by NHS North West London Pathology. A phased transition from the Architect to Alinity platform occurred in 2018 to 2019. The Alinity analysers were validated against the previous Architect analysers in compliance with ISO15189, with comparable performance and unchanged reference ranges. Non-pregnant laboratory references ranges were female follicular: <5 nmol/L; female luteal: >20 nmol/L; serum hCG: <5 IU/L (Seo et al., 2020).

Data was collected from electronic patient records. This included demographic data (age, parity, history of fertility treatment and history of previous TEP), biochemical results (serum P4 and hCG), ultrasound features, and details of initial management (expectant, medical and surgical) and management outcome.

### Statistics

Our analysis compared two cohorts: 1) successful expectant, and 2) combined surgical/medical management. Serum P4, hCG, and maximum ectopic diameter size were compared between groups using Student’s t-test (continuous and parametric data) and Mann–Whitney U test (non-parametric data). These data are presented as means ± standard error of the mean (SEM) or medians ± interquartile range (IQR) as appropriate for the distribution normality. All p-values were two-tailed, and significance defined as p<0.05.

Univariate linear regression was performed to assess the relationship between serum biomarkers (P4, first hCG, peak hCG) and maximum ectopic diameter size, with maximum ectopic diameter as the dependent variable and each biomarker as an independent variable. An additional model evaluated the correlation between P4 and first hCG. The slope of each regression line was tested for significance using an F-test, and regression coefficients with 95% confidence intervals (CI) reported. Model performance was summarised using the coefficient of determination (R²) to indicate the proportion of variance explained.

Multivariate logistic regression analysis was performed to evaluate the combined predictive value of serum biomarkers (P4, peak hCG, first hCG) for successful expectant management. The model was adjusted for age but not IVF or previous TEP as the numbers were small and confined to the combined surgical/medical group. The small number of events would cause model instability. Receiver operating characteristic (ROC) curves were generated for individual biomarkers (P4, first hCG, peak hCG) and for the multivariate model to compare predictive performance. The area under the ROC curve (AUC) with 95% CI was calculated to assess discriminatory accuracy. Comparisons of AUCs were performed using the DeLong method. Internal validation using Harrell’s optimism-corrected bootstrap resampling with 1000 bootstrap samples. Optimal thresholds for predicting successful expectant management were identified from ROC curves (unadjusted serum cut-offs).

A decision tree was developed using recursive partitioning analysis with log_10_ first hCG and P4 as predictors (complexity parameter of 0.01, minimum node size of 10 observations, and maximum tree depth of 3). Model discrimination was assessed by ROC analysis and AUC, with optimism-corrected bootstrap validation as above.

Descriptive statistics and group comparisons were done using GraphPad Prism version 9.0 (GraphPad Software, San Diego, CA, USA). Initial univariate and multivariate regression were conducted in SPSS (version 30, IBM Corp., Armonk, NY, USA). Further regression modelling, ROC, bootstrapping, decision-tree analyses were performed in R (version 4.5.3, R Foundation for Statistical Computing, Vienna, Austria) using Rstudio (version 2026.04.0+526, Posit Software, Boston, MA, USA), and pROC, dplyr, binom, rms, rpart, rpart.plot packages.

### Ethics declarations

The Research Ethics Committee, Cardiff, UK as well as Chelsea and Westminster NHS Foundation Trust, London, UK approved this study (Ref: 25/WA/0271). The study was conducted in accordance with the human research ethics guidelines of the authors’ institution. Informed consent was not required as data was fully de-identified and analysed retrospectively.

## Results

### Demographic and clinical characteristics

In total, N=91 patients were included for analysis. The median age was 32.5 years, the majority were nulliparous, with spontaneous pregnancies and no history of TEP (Table 1). For the whole cohort, the median gestational age at diagnosis of TEP was 42 days (range, 23-74 days) and the mean maximum diameter of ectopic pregnancy was 20mm (range, 6-97 mm). While patient cohorts were not intentionally matched by design, we compared demographic characteristics between each management group (Table 1). There was no significant difference in age (p=0.3958), ethnicity (p=0.3379) or parity (p=0.6993).

**Table 1:**
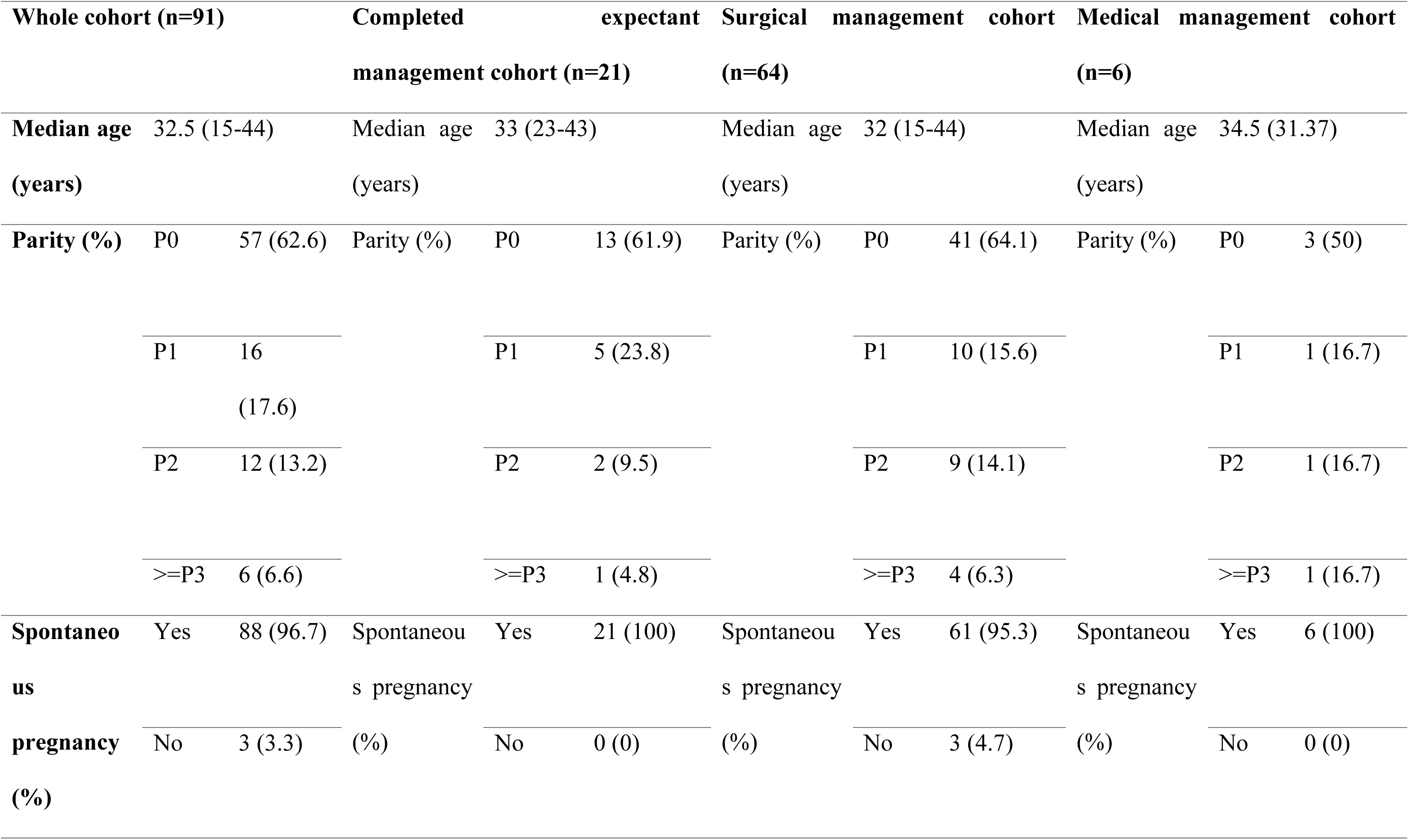
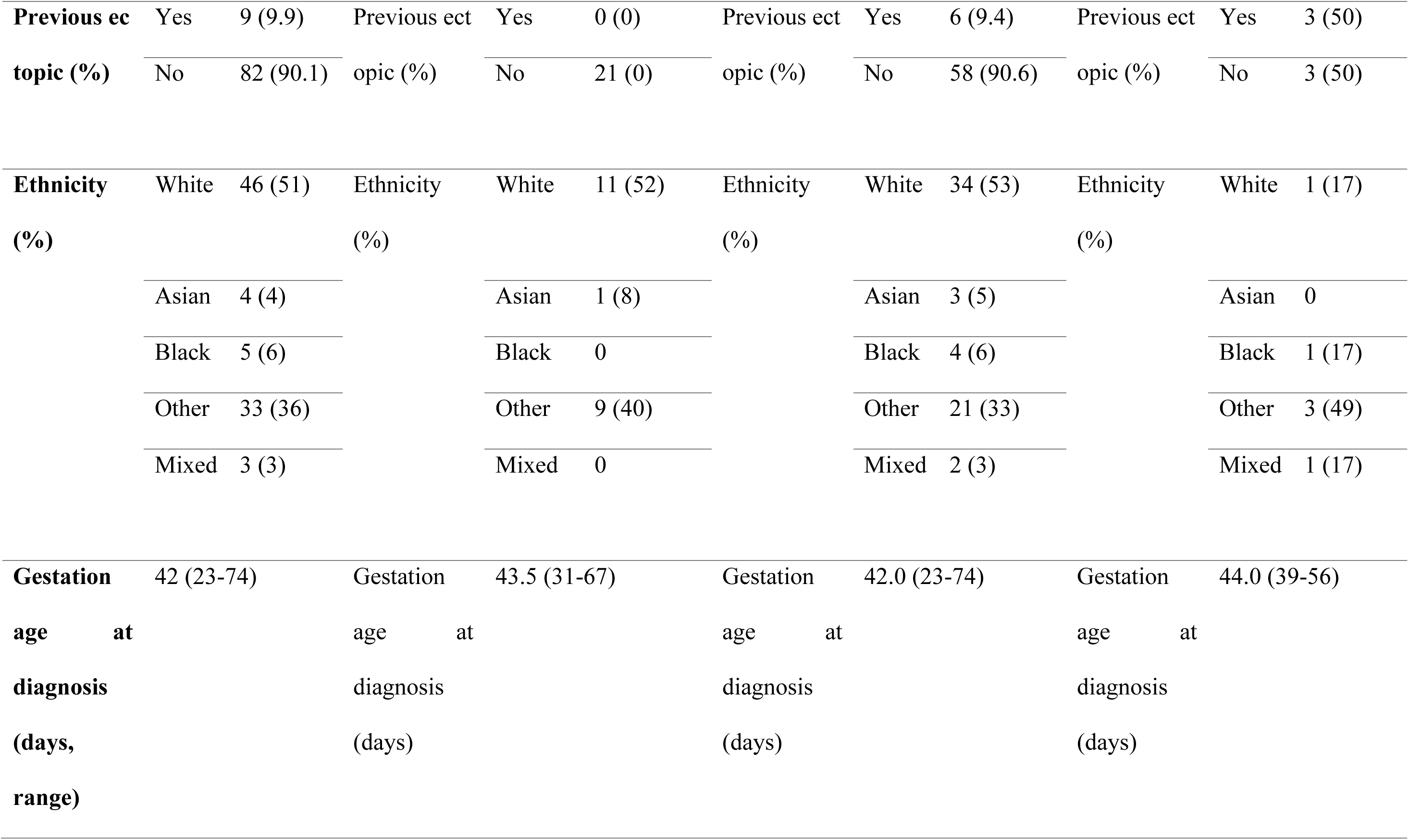
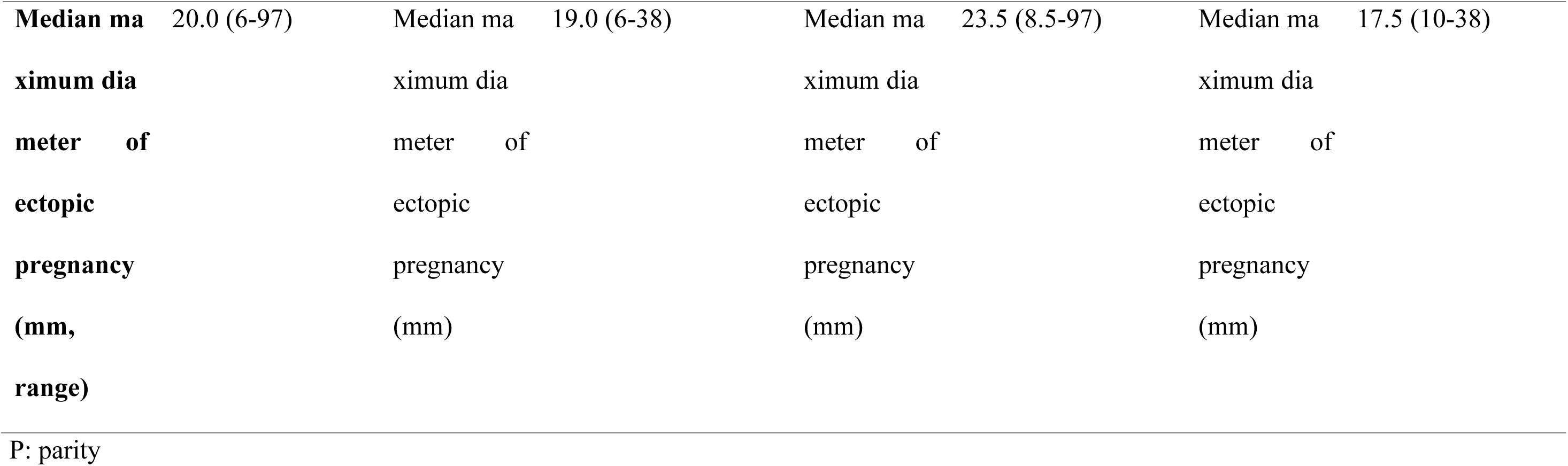
Demographic characteristics and characteristics of ectopic pregnancies in each group.

### Treatment pathways

36% (N=33) of women in the cohort were initially managed expectantly. Follow-up was complete once hCG had returned to pre-pregnancy levels or surgical treatment was required.

At the end of follow-up (median 21 days, range 0-83 days), 23% (n=21) successfully completed expectant management and 77% (n=70) completed surgical/medical management. 64% (n=21/33) of women who were initially managed expectantly successfully completed expectant management. A summary flowchart of patient inclusion and management is shown in Figure 1.

Three women initially offered expectant management fell outside NICE and local eligibility criteria and were therefore excluded from the analysis: two women had an ectopic measuring >35mm (38mm and 54mm, P4 of 36 nmol/L and 9 nmol/L) and one woman with a hCG>1500 (1599 IU/L with a P4 of 7 nmol/L).

### Serum P4 and hCG levels across cohorts

Serum P4 and hCG levels were compared between cohorts. P4 concentrations were significantly lower in the expectant management group (median 3.00, IQR 2.00–8.50) compared with those requiring combined surgical/medical management (median 17.00, IQR 5.75–29.25; U=1125, Z=3.68, p<0.001; Figure 2a). Similarly, first hCG values were significantly lower in those who successfully completed expectant management (median 259 IU/L, IQR 101.50–484.00) compared to those undergoing intervention (median 1269 IU/L, IQR 525–3444; U=1196, Z=4.34, p<0.001; Figure 2b). Peak hCG levels demonstrated a comparable pattern, with lower values observed in the expectant group (median 270 IU/L, IQR 101.50–612.00) compared with the combined surgical/medical group (median 1269 IU/L, IQR 532.00–3444.00; U=1192, Z=4.31, p<0.001; Figure 2c). These univariate findings indicate that lower hCG and P4 concentrations are associated with successful expectant management at an unadjusted level.

**Figure 2:**
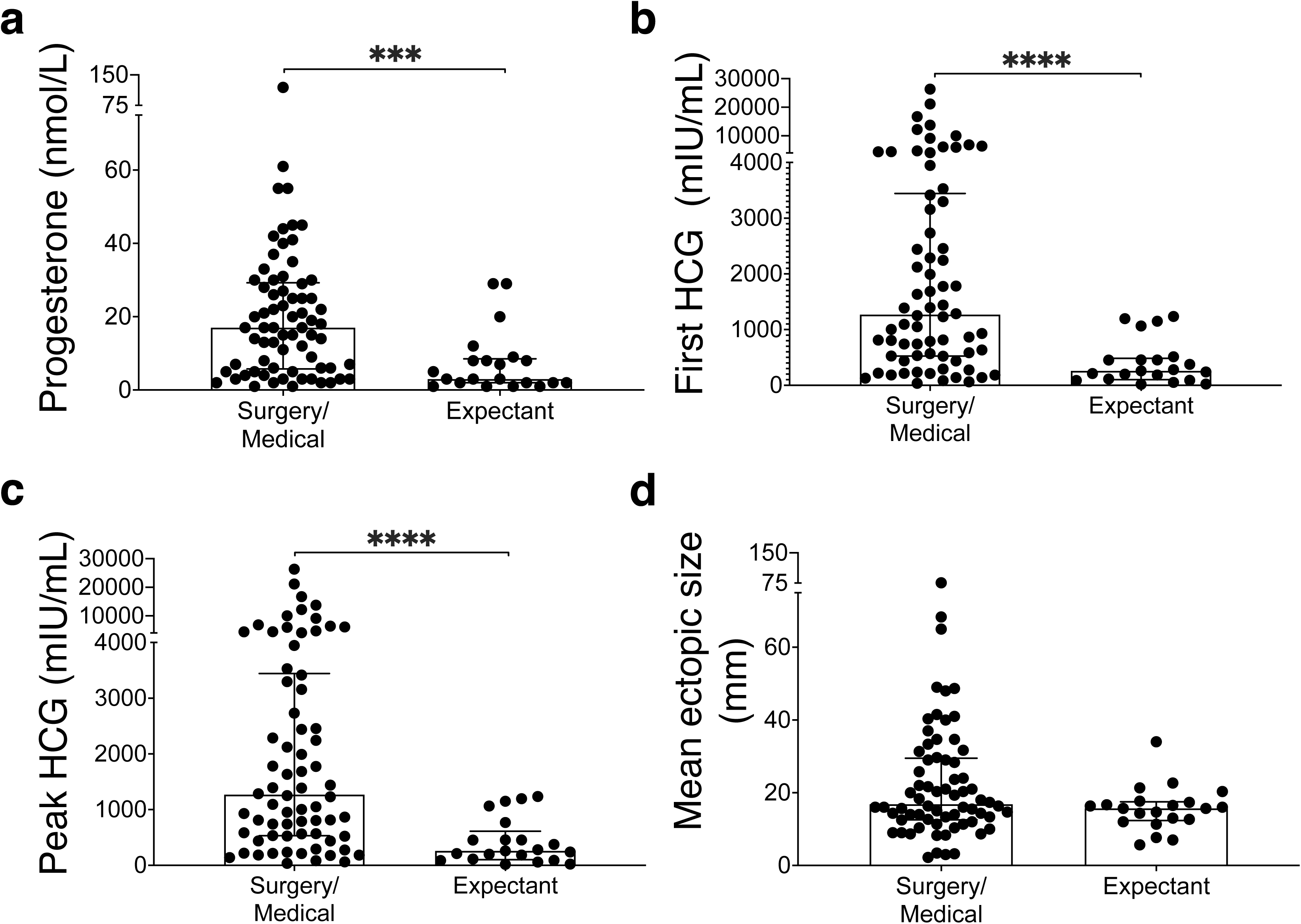
Comparison of P4 and hCG levels and mean maximum ectopic size between expectant management and surgical/medical management combined groups. A) P4, B) First hCG, C) Peak hCG levels, and D) Mean maximum ectopic size was analysed between groups. All data are median with IQR. Statistical analysis was by Mann-Whitney U (for 2 groups). P-values are 2-tailed and defined as *** p<0.001 and **** p<0.0001.

To determine whether these variables independently predicted management outcome, binary logistic regression, adjusted for age, was performed (Table 2). In an initial multivariable model including first hCG, peak hCG, and P4, none of the variables were independently associated with outcome, likely reflecting multicollinearity between first and peak hCG, which are biologically related measures. To address this, hCG variables were log-transformed (log_10_) to reduce skewness and analysed in separate models. In these refined analyses, log_10_ first hCG (p=0.013) and log_10_ peak hCG (p=0.015) were significantly associated with outcome, with higher values suggesting a reduced likelihood of successful expectant management. In contrast, P4 was not independently associated with outcome.

**Table 2:** Multivariate model outputs.

| **Multivariate model of First hCG, Peak hCG and Progesterone** |  |  |  |
| --- | --- | --- | --- |
| **Variable** | **Adjusted OR** | **95% CI** | **p-value** |
| **First hCG** | 1.000 | 0.993-1.007 | 0.987 |
| **Peak hCG** | 1.001 | 0.994-1.009 | 0.745 |
| **Progesterone** | 1.060 | 0.995-1.129 | 0.069 |
| **Multivariate model of Log10 (Peak hCG), Progesterone, and combined (Log10 (peak hCG) and P4)** |  |  |  |
| **Variable** | **Adjusted OR** | **95% CI** | **p-value** |
| **Progesterone** | 0.886 | 0.664-1.183 | 0.411 |
| **Log 10(peak hCG)** | 0.157 | 0.036-0.676 | 0.013 |
| **Combined** | 1.024 | 0.928-1.130 | 0.638 |
| **Multivariate model of Log10 (First hCG), Progesterone, and combined (Log10 (first hCG) and P4)** |  |  |  |
| **Variable** | **Adjusted OR** | **95% CI** | **p-value** |
| **Progesterone** | 0.877 | 0.663-1.161 | 0.359 |
| **Log 10(first hCG)** | 0.157 | 0.037-0.657 | 0.011 |
| **Combined** | 1.028 | 0.935-1.130 | 0.572 |

hCG: human chorionic gonadotrophin; OR: odds ratio; CI: confidence interval

Interaction terms between P4 and first and peak hCG were also explored to assess for effect modification (Table 2). No significant interactions were identified (p=0.598 and p=0.978, respectively), indicating that the relationship between hCG and management outcome does not vary according to P4 levels. Variance inflation factors (VIFs) were also calculated for all age-adjusted regression models to assess multicollinearity between predictors (Supplementary Table 1).

We compared tubal ectopic size between groups given previous research has demonstrated a relationship between serum P4, hCG levels and tubal ectopic size (Guvendag Guven et al., 2006, Ucyigit et al., 2021) and the 35mm NICE threshold for expectant management eligibility (NICE, 2023). Our results showed no difference between the expectant management group and the combined surgical/medical cohort (Figure 2d). On linear regression, first hCG (p=0.027; R²=0.055) and peak hCG (p=0.028; R²=0.054) demonstrated positive correlations with maximum tubal ectopic size, whereas P4 did not (p=0.848; R²=0.00042) (Supplementary Figure 1a-1c). P4 levels correlated positively with first hCG (p=0.019; R²=0.061) (Supplementary Figure 1d).

### Sub-group analysis of Successful versus Unsuccessful Expectant Management

Additional analyses were performed on a cohort of 30 patients (21 successful and 9 unsuccessful expectant outcomes) using the Mann–Whitney U test. P4 was found to differ significantly between successful and unsuccessful expectant management groups (U=48.50, z=−2.093, p=0.036), suggesting a potential association with outcome (Supplementary Figure 2a). In contrast, peak hCG was not significantly different between groups (U=74.00, z=−0.928, p=0.372) (Supplementary Figure 2b). Similarly, log_10_ peak hCG and maternal age showed no association with expectant outcome (U=74.00, z=−0.928, p=0.372; U=108.00, z=0.613, p=0.563 respectively) (Supplementary Figure 2c-2d).

A binary logistic regression model was subsequently constructed to assess whether P4 and log₁₀ peak hCG independently predicted expectant outcome. The overall model was not statistically significant (χ²(2)=4.408, p=0.110), with modest explanatory power (R²=0.194) and an overall classification accuracy of 66.7%. Within this model, neither P4 (odds ratio [OR] 0.93, 95% CI 0.85–1.02, p=0.109) nor log₁₀ peak hCG (OR 0.52, 95% CI 0.06–4.60, p=0.553) were independent predictors of expectant management success.

### Diagnostic accuracy of P4 and hCG

The diagnostic performance of P4 and hCG (age adjusted) in predicting successful expectant management was evaluated using ROC curve analysis. Log_10_ peak hCG demonstrated good discriminative ability (AUC 0.811), as did log_10_ first hCG (AUC 0.814), while P4 demonstrated moderate discrimination (AUC 0.766) (Figure 3A).

**Figure 3:**
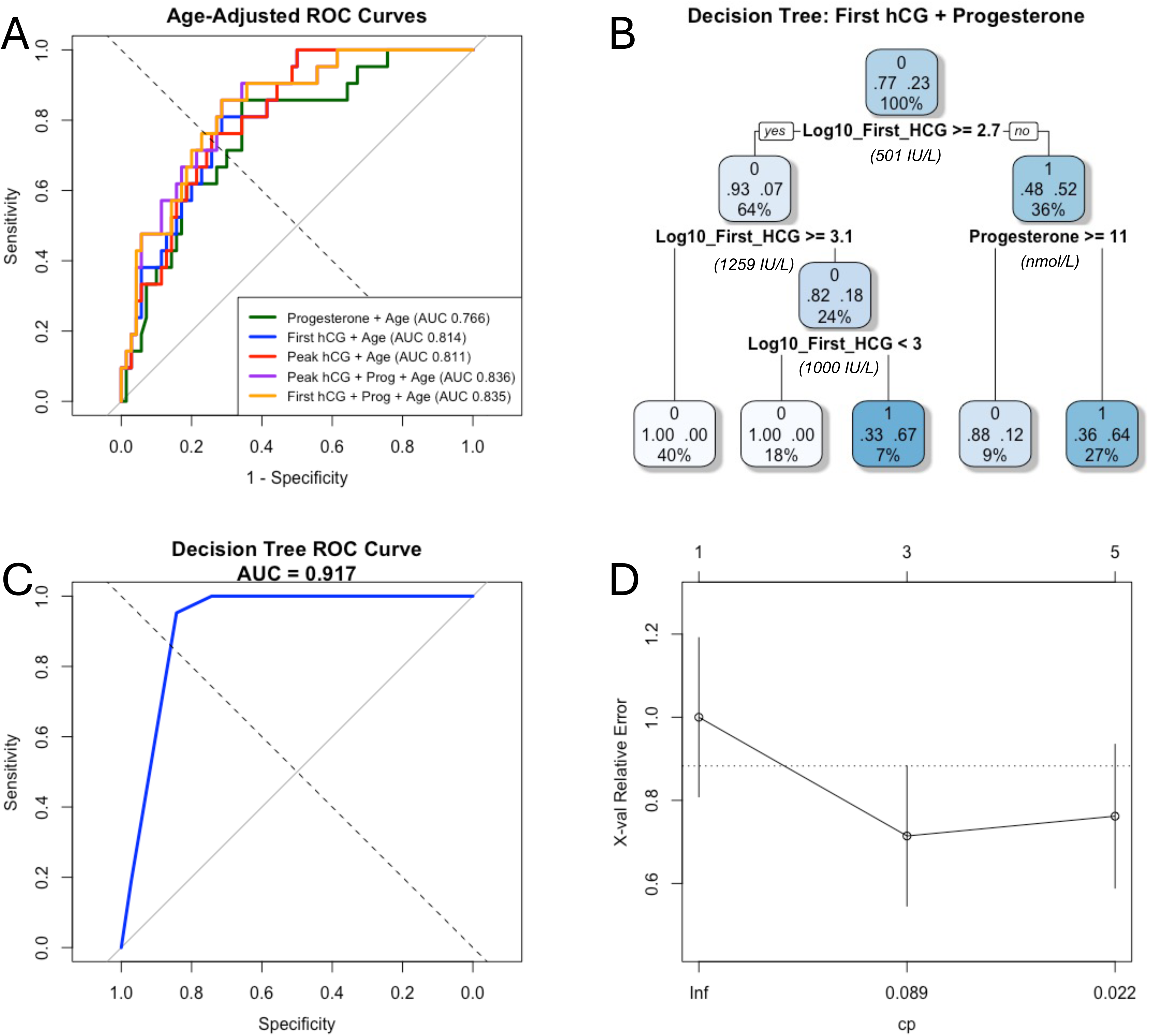
Receiver operating curve (ROC) curve analysis with area under the curve (AUC) values for P4 and hCG, and Decision tree model with first hCG and P4. A) We compared the performance of P4 and hCG at predicting successful expectant management using surgical/medical management cohort as a control group. Our results show A) P4 (green), AUC=0.765 (95% CI=0.652–0.879); log_10_ (first hCG) (blue), AUC=0.814 (95% CI 0.722–0.905); log_10_ (peak hCG) (red), AUC=0.811 (95% CI=0.718–0.903), combined model of log_10_ (peak hCG) and P4 (purple), AUC=0.833 (95% CI=0.739–0.926), and combined model of log_10_ (first hCG) and P4 (yellow), AUC=0.832 (95% CI=0.738–0.926). ROC curve analysis evaluating the performance of P4 and hCG in distinguishing between successful expectant and surgical/medical management. The area under the ROC curve (AUROC) was calculated to quantify overall diagnostic accuracy. An AUROC of 1.0 indicates perfect discrimination, while 0.5 indicates no discriminative ability. B) Decision-tree model branching with the initial split using log_10_ first hCG, followed by P4 thresholds within intermediate-risk groups. Terminal nodes (0=surgical/medical, 1=expectant) display the predicted probability together with the proportion of patients within each node. C) ROC curve for model. D) Root node classification error was 23.1% (21/91). Cross-validation showed that a two-split tree reduced the relative classification error from 1.00 to 0.67, with a corresponding cross-validated error of 0.71. With additional tree complexity beyond four splits, predictive performance did not substantially improve (cross-validated error 0.76). This supports the use of a parsimonious model structure.

Comparison of ROC curves using DeLong’s test showed no significant difference between log_10_ first and peak hCG (p=0.759), indicating equivalent predictive performance. Although first hCG demonstrated a higher AUC than P4, this difference was not statistically significant (p=0.344).

A combined model incorporating log_10_ first hCG and P4 demonstrated a slightly higher AUC (0.835) (Figure 3A), but this was not significantly different from first hCG alone (p=0.372), indicating no meaningful improvement in predictive accuracy with the addition of P4 (Supplementary Table 2). For comparison, log_10_ peak hCG and P4 was also analysed and showed an AUC of 0.836, which was not significantly different to first hCG alone. DeLong’s test for two correlated ROC curves of (age adjusted) log_10_ peak hCG + P4 vs log_10_ first hCG + P4 showed no statistically significant difference between the combined models (Z=0.229, p=0.819, CI −0.010 to 0.013), indicating no evidence of superiority of either model.

Due to the small number of TEP cases, we performed internal validation using Harrell’s optimism-corrected bootstrap resampling with 1000 samples, and chose this rather than split-sample validation, to maximise the efficient use of our available data (Supplementary Table 3, Supplementary Figure 3). Minimal optimism was observed across all models. Optimism-corrected AUCs for P4, log_10_ first hCG and log_10_ peak hCG were 0.747, 0.799, and 0.800, respectively. Combined models log_10_ peak hCG + P4 and log_10_ first hCG + P4, demonstrated the highest discriminatory performance, with AUCs of 0.815 and 0.814 respectively, following optimism correction. Overall, there was minimal optimism (0.011–0.021), indicating minimal evidence of model overfitting.

Optimal thresholds (unadjusted) for each biomarker were derived using the Youden Index to maximise sensitivity and specificity. For log_10_ peak hCG, the optimal cut-off was 2.688, corresponding to approximately 488 IU/L. For log_10_ first hCG, the optimal threshold was 2.713 (approximately 517 IU/L), yielding a sensitivity of 80.9% and specificity of 77.1%. For P4, the optimal cut-off was 12.5, with a sensitivity of 85.7% and specificity of 62.9% (Table 3). Optimum thresholds for both combined models are presented in Table 3.

**Table 3:** Thresholds for predicting successful expectant management of ectopic pregnancy.

|  | **Youden optimal threshold** | **Sensitivity (95%CI)** | **Specificity (95% CI)** |
| --- | --- | --- | --- |
| Progesterone | <12.5 nmol/L | 0.86 (0.48-1.00) | 0.63 (0.21-0.76) |
| First hCG | <517 IU/L | 0.81 (0.38-0.95) | 0.77 (0.44-0.86) |
| Peak hCG | <488 IU/L | 0.90 (0.67-1.00) | 0.79 (0.46-0.89) |
| Combined model of: Log10 Peak hCG and Progesterone | <0.22 | 0.86 (0.57-1.00) | 0.71 (0.34-0.86) |
| Combined model of: Log10 first hCG and Progesterone | <0.26 | 0.81 (0.48–0.95) | 0.77 (0.40–0.89) |

hCG: human chorionic gonadotrophin; OR: odds ratio; CI: confidence interval

### Proposed decision tree using P4 and hCG

Since our work was intended for hypothesis generation, we also decided to explore a different approach and use the data to propose a decision tree model using first hCG followed by P4. At the initial split with a log_10_ first hCG value of 2.7 (approximately 500 IU/L), lower hCG levels were associated with a higher probability of expectant management success (Figure 3B). Among patients with log_10_ first hCG ≥3.1 (approximately 1250 IU/L), the probability of successful expectant management was very low. Among patients with lower first hCG levels, P4 further stratified risk, with levels <11 nmol/L associated with a substantially increased probability successful expectant management (64%). The model demonstrated good discrimination, with an AUC of 0.917 (Figure 3C). However, our dataset is small and decision tree models may be susceptible to instability in smaller datasets. Therefore, internal bootstrap validation was performed to reduce optimism in performance estimates (optimism 0.004). Optimised AUC was 0.913 (95% CI: 0.787-0.978), indicating minimal overfitting.

## Discussion

The focus of our study was to assess whether P4 levels in women with confirmed TEP are associated with their final management outcome. We adopted a pragmatic approach by comparing women who successfully completed expectant management with those who proceeded to require either medical or surgical management (“combined surgical/medical”). This allowed us to explore the clinical utility of P4, hCG and P4 in combination with hCG within a realistic analytical framework for this scoping study. This was in part due to study design constraints, namely a single centre retrospective dataset. Our results show that lower P4 levels at presentation were associated with successful expectant management of TEP. We also found that P4 in addition to hCG provided limited adjunctive value. Overall, hCG was the better performing prognostic biomarker and P4 was not an independent predictor in multivariable models. We also explored a decision tree model with first hCG followed by P4, and this demonstrated the adjunctive potential of P4 in clinical practise. However, our dataset is significantly restricted by small numbers, and a single centre retrospective design that inherently risks clinical bias.

HCG and P4 are essential for early pregnancy, with hCG maintaining corpus luteum function and promoting trophoblast invasion and placentation, while P4 supports endometrial receptivity, immune tolerance, and uterine quiescence (Makrigiannakis et al., 2017, Shah et al., 2019). Although low or declining levels of these hormones are associated with early pregnancy loss, they often reflect underlying non-viability rather than being the primary cause (Deng et al., 2022, Johnson et al., 1993, Wang et al., 2022). Biologically, our findings support the view that hCG and P4 provide complementary information that reflect different physiological processes. Both first and peak hCG showed a weak correlation with tubal ectopic size compared to P4 which showed no such relationship. This has been previously observed in other studies (Elson et al., 2004, Mavrelos et al., 2013, Trio et al., 1995). This dissociation is plausible because P4 in early pregnancy is predominantly determined by corpus luteum function (Duncan, 2021, Ku et al., 2021).

HCG thresholds used for expectant management vary throughout the literature. In a prospective single-centre cohort study, Mavrelos et al. proposed a hCG threshold of <1500 IU/L to identify women suitable for expectant management (Mavrelos et al., 2013). However, as management decisions were based on patient choice rather than randomisation, 19 of 165 eligible women opted for surgery. Of the remaining 146 managed expectantly, 71% successfully completed expectant management. Two women were lost to follow-up, and among the 144 women with complete outcome data, 27% experienced unsuccessful expectant management. Similarly, Trio et al. conducted a prospective case-control study reporting a success rate of 73% managed expectantly using a hCG threshold <1000 IU/L (Trio et al., 1995). Rodrigues et al. performed a single-centre retrospective study without a predefined hCG threshold and reported a 49% success rate (99 of 262 TEPs) with hCGs <2500IU/L (Rodrigues et al., 2012). In a single centre retrospective study, Helmy et al. used a hCG<5000 IU/L threshold for asymptomatic women and reported a success rate of 61% (Helmy et al., 2015). The substantial variation in reported success rates (49–73%) suggests that hCG alone is an imperfect predictor of expectant management success.

Our analysis demonstrated that P4, first and peak hCG distributions were significantly lower in the cohort that completed expectant management. This is concordant with previous findings (Mavrelos et al., 2013, Memtsa et al., 2020, Elson et al., 2004). Higher levels of hCG and P4 in medically and surgically managed TEP likely represents a greater burden of trophoblastic activity and/or P4 stimulation. Falling hCG levels, in particular >15% in 48 hours, are often successfully managed expectantly (Shulman et al., 2023). In contrast, stagnant or rising hCGs are actively managed medically or surgically because they represent biologically more aggressive ectopic pregnancies (Kugelman et al., 2024).

The AUC for log_10_ first hCG and log_10_ peak hCG alone were comparable and both performed better than P4 alone. However, we achieved the highest AUC by combining P4 with log_10_ peak hCG level, generating a higher AUC than P4 or hCG alone (AUC 0.836 versus P4 0.766, log_10_ peak hCG 0.811, log_10_ first hCG 0.814). Our work suggests that P4 could be used to complement hCG in identifying women likely to have successful expectant management of TEP, potentially reducing the number of women who are misclassified as suitable for expectant management by hCG alone (Elson et al., 2004, Mavrelos et al., 2013, Trio et al., 1995, Helmy et al., 2015, Rodrigues et al., 2012). Nevertheless, comparative analysis of our predictive models showed that neither individual biomarkers nor their combination outperformed peak hCG alone.

Elson et al. prospectively studied 179 women with expectant management and surgical subgroups and identified a lower hCG and P4 in expectant management (hCG 246 [99-536] vs 628 [254-1402] IU/L; P4: 10 [6-22] vs 20 [12-31] nmol/L) (Elson et al., 2004). Using these biomarkers alongside sonographic morphology and gestational age, they created a decision tree analysis that can predict successful expectant management. More recently, Graham et al. conducted a single-centre retrospective cohort study (N=798) where only patients that chose to have expectant management were included in the analysis (Graham et al., 2026). Of these 64% (N=512) successfully completed expectant management, with success seen in lower hCG (205 [91-562] vs 910 [515-1457] IU/L) and P4 levels (9 [5-20] vs 23 [12-34] nmol/L). The group compared morphological and biochemical differences between the two groups to define parameters that could be used in a clinical decision tree. Their decision tree first stratified cases by initial hCG level and further subdivided cases by either P4 (<7.0ng/ml [23.8nmol/L]) or average ectopic diameter (>16mm). Of note, thresholds were derived from the CHAID algorithm rather than from ROC-based Youden optimisation. Our findings complement these studies. Using ROC analyses of simplified combined hCG and P4 models, we found that combined markers improved specificity compared with P4 alone, but reduced sensitivity, and did not significantly outperform hCG alone. This suggests limited stand-alone clinical value for P4. Since our work was exploratory and intended for hypothesis generation, we used the same approach as Graham et al and Elson et al and developed a decision tree model with sequential use of first hCG followed by P4. Surprisingly, the use of both markers was associated with an improved probability of successful expectant management (AUC 0.917). However, we acknowledge that the referenced studies have a significantly greater sample size, greater methodological robustness and clearer clinical applicability.

Overall, our proposed optimised cut-offs of <488 IU/L for peak hCG and <12.5 nmol/L for P4 are broadly consistent with those reported by Graham et al. and align with thresholds from studies on expectant management of pregnancies of unknown location (Cordina et al., 2011, Day et al., 2009, Elson et al., 2004, Graham et al., 2026). For example, Cordina et al. and Corsan et al. reported P4 thresholds of <10 ng/mL (<31.8 nmol/L) and <7.0 ng/mL (<22.2 nmol/L), respectively, in cohorts where some pregnancies ultimately resolved spontaneously, including ectopic pregnancies (Cordina et al., 2011, Corsan et al., 1995). Importantly, in our cohort, the Youden optimised threshold for P4 demonstrated a high sensitivity (0.86) but with moderate specificity (0.63), indicating a good ability to identify individuals likely to experience a successful expectant management, albeit with a higher rate of false positives.

This study’s strengths include the use of consecutive cases from a real-world EPAU service, ultrasound-confirmed diagnosis, and standardised laboratory assays for P4 and hCG. Our data adds to the existing literature by exploring the potential of P4 and hCG to determine success of expectant management using different approaches. However, our study has several significant limitations. Firstly, this is a small study in a single centre, in which we compared the cohort of women who successfully completed expectant management to the whole cohort of women presenting with TEP making it significantly underpowered to assess the prognostic value of hCG and P4 as biomarkers. Whilst this may be an appropriate design for an initial scoping study to generate research questions and hypotheses, appropriately powered and substantially more robust study designs are needed to address the clinical issue. Crucially, our decision to compare outcomes between expectant management and combined surgical/medical management is unable to accurately assess expectant management success without cohorts of successful versus failed expectant management. Despite attempting to address this with a subgroup analysis, it remains severely underpowered. In addition, all included patients had already been clinically selected for expectant management before inclusion. As such we acknowledge that some cases managed medically or surgically may have been biologically suitable for successful expectant management, which cannot be fully accounted for in the retrospective design. Moreover, because we applied the NICE inclusion criteria in our study, higher-risk cases were systematically removed from our sample, introducing further selection bias that most likely led to our lower threshold values compared to studies by Ransom et al. and Corsan et al. (Ransom et al., 1994, Corsan et al., 1995). Other factors that may influence adoption of expectant care include clinician and patient anxiety, longer follow-up care versus surgical or medical options, and concerns regarding future fertility. These were not addressed in our work but should be considered in future research.

In conclusion, our findings suggest that a lower serum P4 is associated with successful expectant management of TEP. However, neither P4 alone nor the addition of P4 to hCG clearly outperforms hCG-based models. Decision tree analysis suggests that P4 may have a role as an adjunctive marker but better designed prospective studies using hCG and P4 as triaging markers are needed to explore this further.

## Acknowledgements

The authors thank the patients and clinical staff at Chelsea & Westminster Hospital who participated in this study.

## Author contributions

DO and NS had a substantial contribution to the conception and design. AKA, MP, DO, KL, NS, AA, and AN were responsible for acquisition, analysis, and interpretation of data. AKA, MP, NS and DO all drafted the manuscript. All authors contributed to revisions of the manuscript. All authors meet established authorship criteria (ICMJE), have approved the final version, and accept accountability for the integrity of the work.

## Data availability statement

The datasets generated and analysed during the current study are derived from de-identified clinical data and are not publicly available due to ethical and privacy restrictions. However, they may be made available from the corresponding author on reasonable request for non-commercial research and with appropriate institutional approvals.

