## Supplementary tables and figures for "Progesterone and hCG in expectant management success in tubal ectopic pregnancy: retrospective single-centre cohort study"

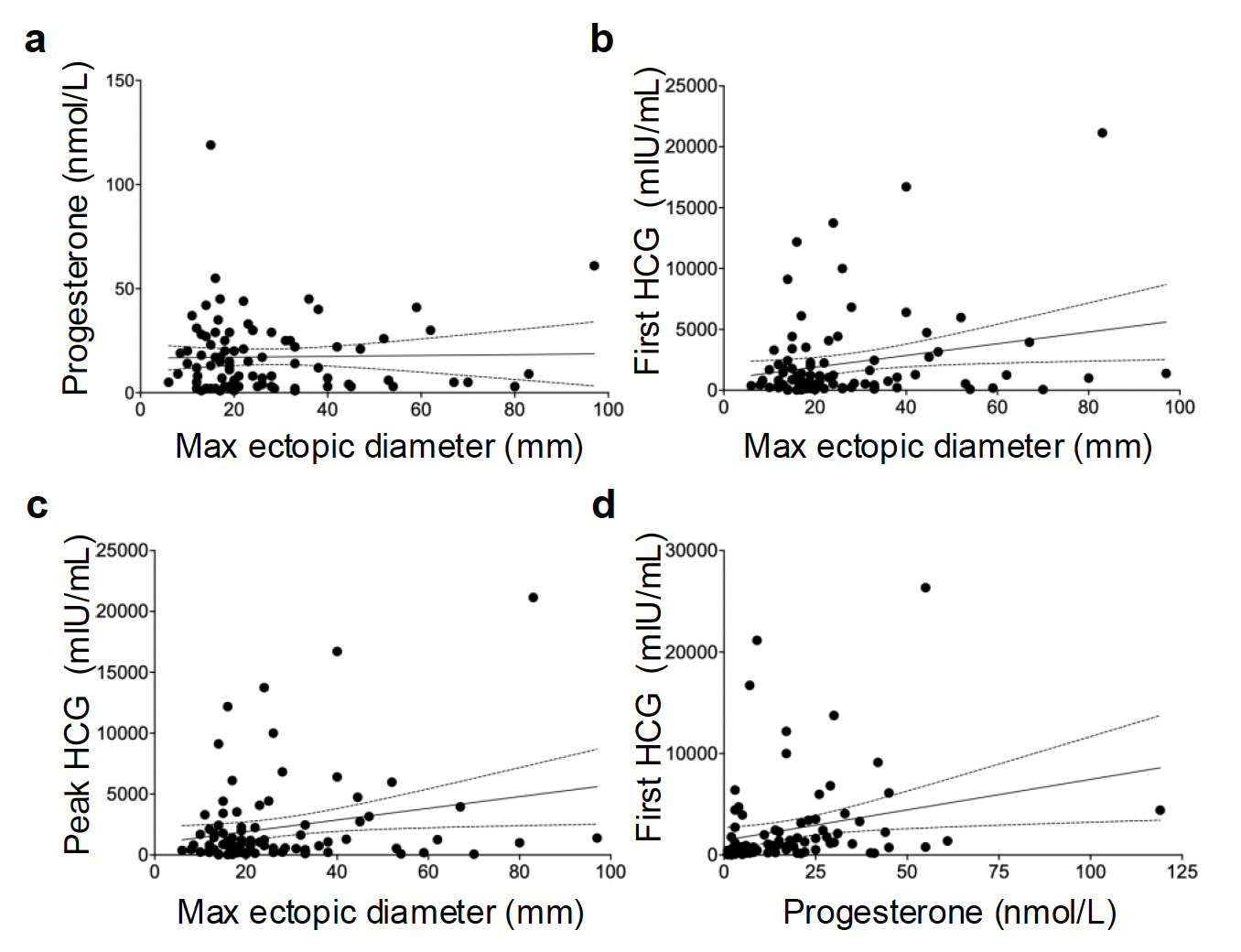

**Supplementary Figure 1: Correlation between P4 (progesterone)/hCG (human chorionic gonadotrophin), and Maximum Ectopic Diameter, as well as between P4 and hCG.** A) P4 and maximum diameter of the ectopic size where the slope was not significant (p=0.848; 95% CI: –0.1919 to 0.2329), with minimal explained variance (R²=0.00042); B) first hCG and maximum diameter of ectopic where the slope was significant (p=0.027; 95% CI: 5.485 to 90.80), with modest explanatory power (R²=0.055); C) peak hCG and maximum diameter of ectopic where the slope was significant (p=0.028; 95% CI: 5.369 to 90.55), with modest explained variance (R²=0.054); and D) P4 and first hCG where the slope was significant (p=0.019; 95% CI: 10.36 to 110.5), with low explained variance (R²=0.061). A simple linear regression model was fitted for each panel, and the significance of the slope was assessed using an F-test. The regression equation, p-value, and coefficient of determination (R²) are reported for each. Dotted lines represent the 95% confidence intervals (CI) of the fitted regression lines.

**Supplementary Table 1: Multicollinearity Diagnostics (Variance Inflation Factors) for Age-Adjusted Logistic Regression Models Predicting Expectant Management Success**

|  | **Model** | **Variable** | **VIF** |
| --- | --- | --- | --- |
| **P4** | P4 + Age | P4 | 1.00 |
| **Age** | P4 + Age | Age | 1.00 |
| **Log10_First_HCG** | Log10 First hCG + Age | Log10_First_HCG | 1.00 |
| **Age** | Log10 First hCG + Age | Age | 1.00 |
| **Log10_Peak_HCG** | Log10 Peak hCG + Age | Log10_Peak_HCG | 1.00 |
| **Age** | Log10 Peak hCG + Age | Age | 1.00 |
| **Log10_Peak_HCG** | Peak hCG + P4 + Age | Log10_Peak_HCG | 1.09 |
| **P4** | Peak hCG + P4 + Age | P4 | 1.09 |
| **Age** | Peak hCG + P4 + Age | Age | 1.00 |
| **Log10_First_HCG** | First hCG + P4 + Age | Log10_First_HCG | 1.10 |
| **P4** | First hCG + P4 + Age | P4 | 1.10 |
| **Age** | First hCG + P4 + Age | Age | 1.00 |

VIF= Variance Inflation Factors; P4=progesterone

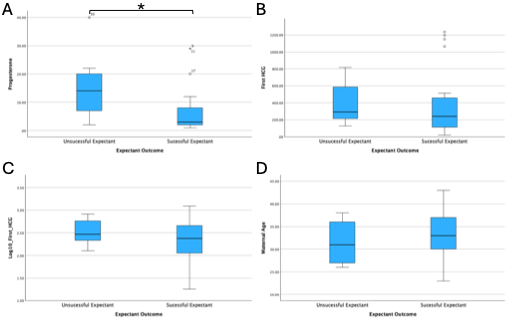
**Supplementary Figure 2: Comparison of P4 (progesterone) and hCG (human chorionic gonadotrophin) levels between unsuccessful and successful expectant management cohorts**. A) P4, B) First hCG, C) Log10 first hCG, and D) Age. All data are median with IQR (interquartile range). Statistical analysis was by Mann-Whitney U test (for 2 groups). P-values are 2-tailed and defined as *p<0.05.

**Supplementary Table 2: Area Under the Curve (AUC) with 95% Confidence Intervals and p-values for individual and combined predictors of Management (age adjusted), including comparisons versus Log10 First HCG**

| Model | AUC | 95% CI | p-value (AUC ≠ 0.5, Z-test) | Comparison (DeLong p-value) |
| --- | --- | --- | --- | --- |
| Log10 First hCG | 0.814 | (0.722–0.905) | <0.001 | Reference |
| Log10 Peak hCG | 0.811 | (0.719–0.903) | <0.001 | 0.759 vs Log 10(peak HCG) |
| P4 | 0.766 | (0.654–0.878) | <0.001 | 0.344 vs Log 10(peak HCG) |
| Log10 peak hCG + P4 | 0.836 | (0.745–0.927) | <0.001 | 0.354 vs Log 10(peak HCG) |
| Log10 first hCG + P4 | 0.835 | (0.743–0.926) | <0.001 | 0.361 vs Log 10(peak HCG) |

P4=progesterone

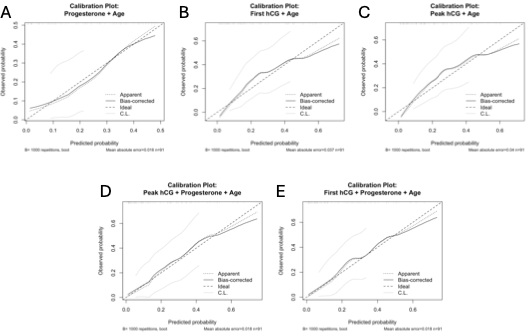

**Supplementary Figure 3: Calibration plots for combined models following bootstrapping.** A) Progesterone, B) First hCG, C) Peak hCG, D) Peak hCG and Progesterone, and E) First hCG and Progesterone. Calibration assessed using Harrell’s optimism-corrected bootstrap resampling with 1000 samples. The diagonal reference line represents perfect agreement between predicted and observed probabilities, while the solid calibration curve represents model performance after bootstrap correction for optimism. Models demonstrated good agreement between predicted and observed outcomes.

**Supplementary Table 3: Area Under the Curve (AUC) with Optimism Corrected AUC following Internal Validation with Bootstrapping**

| **Model** | **Apparent AUC** | **Optimism** | **Optimism Corrected AUC** |
| --- | --- | --- | --- |
| **P4** | 0.766 | -0.019 | 0.747 |
| **Log10 First hCG** | 0.814 | -0.015 | 0.799 |
| **Log10 Peak hCG** | 0.811 | -0.011 | 0.800 |
| **Log10 Peak hCG +P4** | 0.836 | -0.021 | 0.815 |
| **Log10 First hCG +P4** | 0.835 | -0.021 | 0.814 |

 P4=progesterone
